# A Sleep Disorder Detection Model based on EEG Cross-Frequency Coupling and Random Forest

**DOI:** 10.1101/2020.06.10.20126268

**Authors:** Stavros I. Dimitriadis, Christos I. Salis, Dimitris Liparas

## Abstract

Sleep disorders are medical disorders of a subject’s sleep architecture and based on their severity, they can interfere with mental, emotional and physical functioning. The most common ones are insomnia, narcolepsy, sleep apnea, bruxism, etc. There is an increased risk of developing sleep disorders in elderly like insomnia, periodic leg movements, rapid eye movement (REM) behaviour disorders, sleep disorder breathing, etc. Consequently, their accurate diagnosis and classification are important steps towards an early stage treatment that could save the life of a patient. The Electroencephalographic (EEG) signal is the most sensitive and important biosignal, which is able to capture the brain sleep activity that is sensitive to sleep. In this study, we attempt to analyse EEG sleep activity via complementary cross-frequency coupling (CFC) estimates, which further feed a classifier, aiming to discriminate sleep disorders. We adopted an open EEG Database with recordings that were grouped into seven sleep disorders and a healthy control. The EEG brain activity from common sensors has been analysed with two basic types of cross-frequency coupling (CFC). Finally, a Random Forest (RF) classification model was built on CFC patterns, which were extracted from non-cyclic alternating pattern (CAP) epochs. Our RF^CFC^ model achieved a 74% multiclass accuracy. Both types of CFC, phase-to-amplitude (PAC) and amplitude-amplitude coupling (AAC) patterns contribute to the accuracy of the RF model, thus supporting their complementary information. CFC patterns, in conjunction with the RF classifier proved a valuable biomarker for the classification of sleep disorders.

## 1. Introduction

Electroencephalography (EEG) is the basic modality in sleep research, providing a non-invasive way to study brain dynamics in relationship to sleep [1]. Adult sleep is composed of non-rapid-eye-movement (NREM) and rapid-eye-movement (REM) brain states, which alternate almost every 90 minutes. Additionally, the NREM state is divided into four sleep stages, NREM1-4 [2]. Sleep is regulated by the circadian rhythm, which is an internal timer mechanism that maintains regular sleep timetable, hormone secretion and sustained oscillations in body temperature [3]. Normal sleep runs for four or five cycles, including a passing from the four NREM stages and the state REM sleep.

Sleep disorders, no matter what the cause is, affect fifty to seventy million Americans spanning all ages and socioeconomic classes [4]. This tremendous number of affected population can be characterized as a public healthy epidemic. This may be primarily due to an intrinsic problem that affects the sleep – wake cycle or secondary due to a medical condition [5]. Primary sleep disorders are divided into dyssomnias, which are further subdivided into intrinsic, extrinsic and circadian or parasomnias, which are subdivided into arousal disorders, sleep-wake transition disorders, parasomnias associated with REM Sleep and a fourth category of parasomnias [6].

The definition and distinction of those sleep stages are based upon the estimation of rhythmic neural oscillations during sleep that demonstrate different patterns [7] and provide a substrate to study how brain rhythms are related to cortical activity patterns and how this correlation varies across arousal levels [8]. Advanced signal processing techniques support a more detailed exploration of brain oscillations during sleep, which further advances our understanding of the related overcoming limitations of traditional quantification approaches, like how many minutes a person spent in each sleep stage, percentage of REM sleep compared to the total sleep time, or ratio-based sleep variables. Detection of sleep oscillations can be realized under visual inspection of oscillation events [9], automatic event detector algorithms like K-complexes [10], spectral analysis techniques [11-12] and cross-frequency coupling (CFC) estimates [13].

Interactions between activities with different frequency content and time scales have been widely explored in neuroscience to define a physiological biomarker, e.g. mild cognitive impairment [14], in mild traumatic brain injury [15], in sleep [13,16], etc. Neural activities with different frequency content have origin in cell population of different size and spatial orientation, while their interactions, named (CFC), have been widely explored over the last years [8,17]. CFC refers to interactions that can occur between brain rhythms with different frequency content, between different signals or within the same signals that capture brain activity of brain areas, respectively [17, 18]. CFC has been adopted in studies that attempt to explore how information propagates across brain areas indicative of distinct neurophysiological functions [18-22]. In general, three basic types of CFC exist: phase synchronization (phase-phase CFC [23-24]), phase-amplitude coupling (PAC) [14,15,21,22] and amplitude-amplitude coupling (AAC) [13,25].

In the current study, we adopt our framework first presented in a single-sensor automatic sleep stage classification with high accuracy based on various types and estimators of CFCs (Dimitriadis et al., 2018). We aim to develop the first model for an accurate sleep disorder detection, based on EEG sleep activity with the incorporation of two types of CFC, the PAC and AAC. We hypothesize that the strength of CFC will be sensitive in different sleep stages across sleep disorders. As a proper classifier, we employ Random Forest (RF), which has proved efficient in many clinical problems [26]. Our first attempt focuses on non-cyclic alternating pattern (CAP) patterns.

## 2. Method

### 2.1 Material

For the purpose of our study, we downloaded all-night polysomnography (PSG) recordings from the well-known database: “The CAP Sleep Database” of “PhysioBank” (https://physionet.org/physiobank/database/capslpdb/) [27, 28]. This study is a collection of 108 PSG recordings that include seven groups with sleep disorders and a normal healthy one that did not present any medical, neurological or psychiatric disorder. The approval of the study has been registered in the Sleep Disorders Center of the Ospedale Maggiore of Parma, Italy [27, 28]. In addition, the clinical information for the whole cohort can be found in the excel file called “gender-age.xlsx”. Well-trained sleep expert neurologists manually scored the sleep recordings based on the Rechtschaffen & Kales rules, classifying epochs as sleep stages 1–4, wake, REM sleep and movement artefacts [29]. Cycling Alternating Pattern (CAP) was determined based on the Terzano rules (phase-A subtypes include A1, A2 and A3) [27].

In our study, we analysed non-CAP epochs by removing epochs related to CAP. An epoch is characterized as either a CAP sequence or a non-CAP. A CAP sequence is composed of a succession of CAP cycles. A CAP cycle is composed of a phase A and the following phase B (Fig. 1). All CAP sequences begin with a phase A and end with a phase B. Each phase of CAP is 2–60 s in duration. The absence of CAP for 60 s is scored as non-CAP. An isolated phase A, (that is, preceded or followed by another phase A but separated by more than 60 s), is classified as non-CAP. The phase A that terminates a CAP sequence is counted as non-CAP. This transitional phase A bridges the CAP sequence to non-CAP.

**Figure 1.**
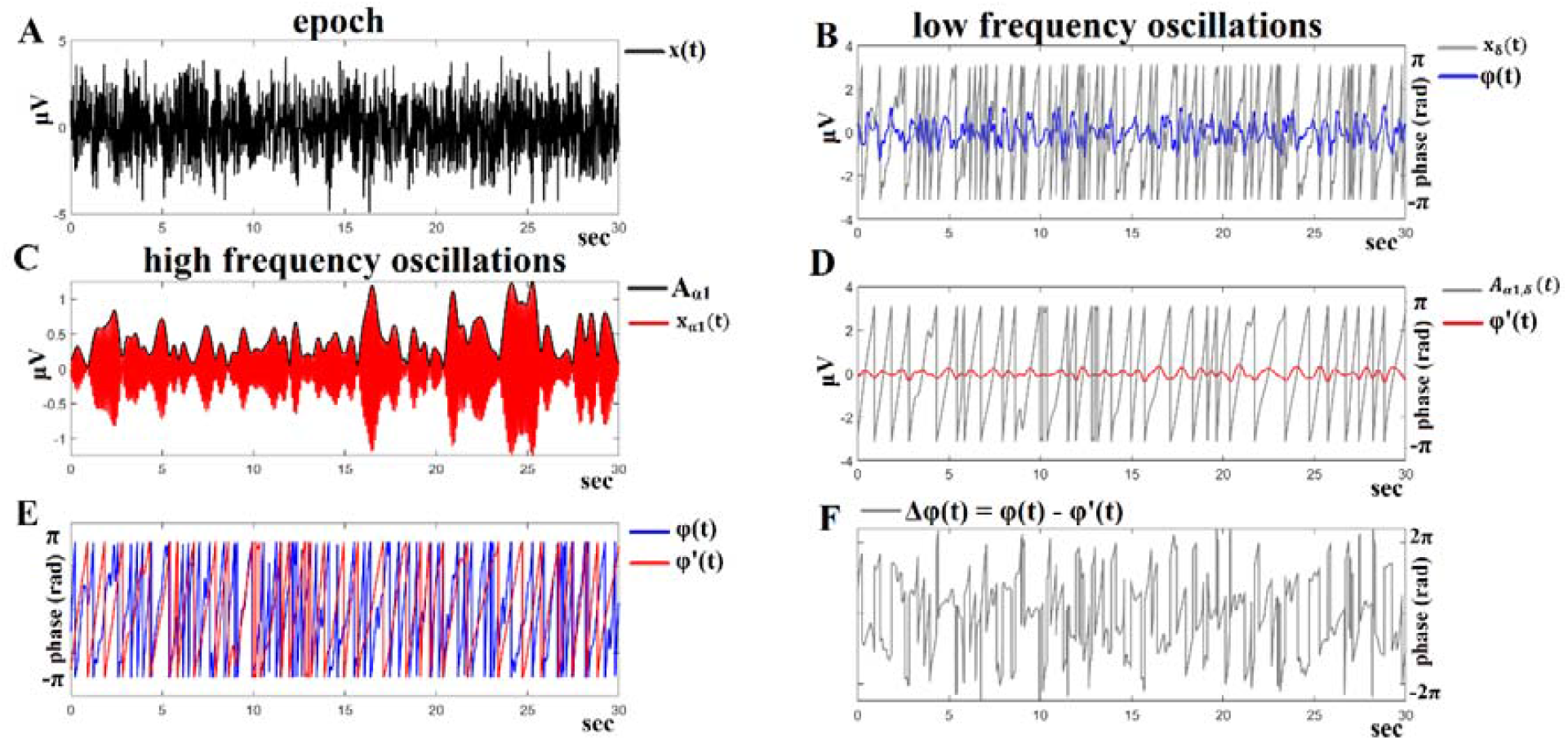
Algorithmic steps for PAC estimation. Using an epoch of 30 sec. length recorded at the P4-O2 sensor from the first subject with periodic leg movements during the NREM3 stage (A), we showed the phase-to-amplitude coupling between δ and α_1_ rhythms. The time series presented in (A) was firstly bandpass filtered using a zero-phase-order filter into a low-frequency δ (0.5–4DHz) component, where its envelope via Hilbert transform was also extracted (B), and secondly into a high-frequency α_1_ (8–13DHz) component, where its envelope was extracted via Hilbert transform (C). We then filtered the amplitude of the envelope high-frequency α_1_ (D) within the δ frequency range (0.5–4DHz). This algorithmic step is important because it will provide us with the δ modulation within the lower α_1_ amplitude. (E) Subsequently, we performed the Hilbert transformation at both the δ-filtered signal and the δ-filtered within the lower-α amplitude, extracting the related phase dynamics and, finally, their phase consistency with iPLV. The phase differences of those two-phase time series are illustrated in (F) and will then enter into the iPLV estimator to quantify the strength of PAC coupling between δ and α_1_ rhythms. Consequently, the aforementioned CFC-PAC analysis will determine how the phase of the lower-frequency component modulates the amplitude of the high-amplitude component.

Furthermore, it should be noted that the EEG recordings were sampled at 100 Hz, 256 or 512 Hz. The waveforms (contained in the .edf files of the database) include at least 3 EEG channels (F3 or F4, C3 or C4 and O1 or O2, referred to A1 or A2), EOG (2 channels), EMG of the submentalis muscle, bilateral anterior tibial EMG, respiration signals (airflow, abdominal and thoracic effort and SaO2) and EKG. Additional traces include EEG bipolar traces, according to the 10-20 international system (Fp1-F3, F3-C3, C3-P3, P3-O1 and/or Fp2-F4, F4-C4, C4-P4, P4-O2).

We selected subjects with the same setup of EEG sensors and with frequency sampling of either 256 or 512 Hz, considering that it is an important parameter in signal processing analysis. In our work, we excluded epochs annotated as movement artefacts. Table 1 summarizes the number of subjects per sleep disorder with their gender distribution and mean age.

**Table 1.** Demographics of the Sleep Disorder Dataset

| Sleep Disorder | No of Subjects | Males | Females |
| --- | --- | --- | --- |
| Bruxism (BRUX) | 2 | 2 (28.50±7.77) |  |
| Sleep-disordered breathing (SBD) | 3 | 4 (71.25±7.22) |  |
| Insomnia (INS) | 9 | 4 (68.00±11.88) | 5 (55.20±4.96) |
| Narcolepsy (NARCO) | 5 | 2 (33.50±13.43) | 3 (30.33±13.05) |
| Nocturnal frontal lobe epilepsy (NFLE) | 40 | 21 (27.80±8.27) | 19 (33.00±12.87) |
| Periodic leg movements (PLM) | 10 | 7 (56.71±7.54) | 3 (51.33±1.15) |
| REM behavior disorder (RBD) | 22 | 19 (70.10±6.65) | 3 (74.66±1.52) |
| No pathology (N) | 16 | 7 (30.42±3.73) | 9 (33.55±6.52) |

### 2.2 Signal Processing

To reduce the effects of artefacts, we performed a combination of Fast Independent Component Analysis (ICA) and maximum overlap discrete wavelet transform (MODWT). Fast ICA was applied to the whole set of EEG sensors, giving an equal number of independent components with a characteristic topology and time course. Traditional artefact correction analysis based on ICA classifies an independent component as artefact or not. In the former case, researchers zeroed the whole component and the corresponding time course was considered as artefact. However, in most studies, an independent component doesn’t reflect an artefact in the whole-time course. As a result, setting it equal to zero leads to the elimination of the true brain activity. For that reason, every independent component time series was decomposed via wavelet transform and only subcomponents related to an artefact were deleted. Finally, the rest of wavelets subcomponents were reconstructed, giving rise to a clean independent component time series, which is then projected back to the original space. We adopted maximal overlap discrete wavelet transform to decompose every time source related to an independent component and especially Daubechies wavelets. We followed this wavelet-driven decomposition every 5 sec and by employing a semi-supervised technique of estimating entropy, kurtosis and skewness, we assisted in the process of zeroing (or not) wavelet subcomponents. We collected entropy, kurtosis and skewness across epochs of 5 sec and sensor time series independently per subject and then we estimated the zscore for each of the three second order statistics. A zscore > 2 at least in one of the three adopted statistics is an indication of an artefactual epoch. We visually inspected every case to further evaluate this classification. Then, cleaned wavelet subcomponents were reconstructed to the original time series. The cleaned time courses of the independent component were projected back to the original recording EEG space. The whole approach has been repeated independently on the studied frequency bands.

Finally, we downsampled the PSG recordings with a frequency sampling rate of 512 Hz or 256 Hz, in order to meet the lowest frequency sampling in the cohort, while subjects with 100 Hz were excluded from further analysis. We selected the following common EEG sensors provided by the cohort as a common template: P4-O2, F4-C4, C4-P4 (three bipolar) and C4-A1 (one monopolar). The number of participants that were used in our study with the related sleep disorder are provided in Table 2.

**Table 2.** A summarization of the indexes of the selected subjects per group.

| **Bruxism** | **Insomnia** | **Healthy Controls** | **Narcolepsy** | **Nocturnal frontal lobe epilepsy** | **Periodic leg movements** | **REM behavior disorder** | **Sleep-disordered breathing** |
| --- | --- | --- | --- | --- | --- | --- | --- |
| brux1, brux2 | isn1-ins9 | n1-n5, n10, n11, n16 | narco1-narco5 | nfle1-nfle40 | plm1-plm10 | rbd1-rbd22 | sdb1-sdb4 |

Scores are provided by experts with the following annotations:

Sleep stage (W=wake, S1-S4=sleep stages, R=REM, MT=body movements). In our study, we focused on S1-S4 and REM sleep stages.

PSD recordings were bandpass filtered in the following seven frequency bands : δ {0.5– 4□Hz}, θ {4–8□Hz}, α_1_ {8–10□Hz}, α_2_ {10–13□Hz}, β_1_ {14–20□Hz}, β_2_ {21–30□Hz} and γ_1_ {31–45□Hz}.

### 2.3 CFC Estimations

#### 2.3.1 Phase-to-amplitude coupling cross-frequency coupling (PAC)

The most studied CFC in the literature is phase-to-amplitude coupling (PAC) [30]. In most studies, PAC was estimated between the phase of the slower rhythm and the amplitude of the faster oscillation. Here, we followed a different analytic approach, which was first presented in our previous works including an EEG sleep study [13, 14].

The PAC analytic approach for a single EEG P4-O2 sensor is described below:

Let x(*i*_sensor_, *t*) be the EEG time series acquired at the *i*_sensor_ -th recording site, and *t*□ = □1, 2, … *T* denotes the sample points. Given a band-passed filtered signal of x(*i*_sensor_, *t*), PAC is estimated between the phase of the lower-frequency (LF) oscillations and the amplitude of the higher-frequency (HF) ones. The following equations give a full representation of the complex transformations of both (LF) *z*_LF_ (t) and (HF) oscillations *z*_HF_ (t) extracted via the Hilbert transform (HT[.]).

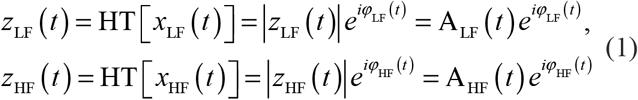

This step gives rise to two time series that capture the envelope A_HF_ (*t*) and the instantaneous phase □_LF_ (*t*) dynamics. The next step involves the band-pass filtering of the higher frequency oscillations A_HF_ (*t*) within the range of LF oscillations and the resulting signal is Hilbert-transformed to extract its phase-dynamics component φ′_LF-HF_ (t):

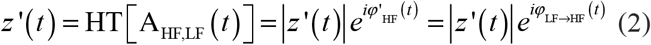

The aforementioned equation reflects the HF-oscillations amplitude modulated by the phase of the LF-oscillations. The resulting time series are employed to estimate PAC, by means of phase-locking (or synchronization index) technique.

**Phase locking value (PLV)** is defined as follows:

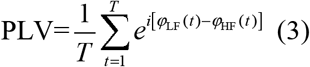

Imaginary part of PLV (iPLV) is defined as follows:

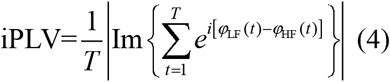

iPLV ranges between 0 and 1, with higher values indicating stronger comodulating PAC interactions.

Figure 1 illustrates every step of the cross-frequency coupling (CFC) – phase-to-amplitude coupling (PAC) analysis, where a time series of 30 sec. length was adopted from the first subject with periodic leg movements during the NREM3 stage at the P4-O2 sensor. PAC estimations are examined between the LF and HF oscillations, which corresponded to δ and α_1_ brain rhythms, respectively. The original time series is presented in Figure 1A. The LF version of the original signal, along with the trace of its instantaneous phases □_δ_(*t*) are depicted in Figure 1B. The corresponding HF version of the initial signal, along with the trace of its envelope A_α1_ are depicted in Figure 1C. Figure 1D shows the low-pass filtered, within δ frequency range, version of the previous envelope [i.e., the A_α1_, δ(*t*) signal]. The saw-like trace corresponds to its instantaneous phases □′_α1_(*t*). The □_δ_(*t*) and □‘_α1_(*t*) traces have been plotted aligned in Figure 1E, forming the resulting instantaneous phase differences, as shown in Figure 1F. This resulting sequence, representing phase-differences Δ □ (*t*), enters in equation (2) and will be integrated across time by averaging directional vectors e^*i*Δ□(*t*)^ in the complex domain. The length of the original time series should be long enough, so that the iPLV index results into a reliable estimation of PAC.

In the present study, we estimated PAC for a total of (7□x□6)/2□ = □21 cross-frequency pairs per epoch.

#### 2.3.2 Amplitude-to-amplitude cross-frequency coupling (AAC)

AAC is estimated between the Hilbert envelopes of band-pass filtered time series and by adopting a correlation coefficient to quantify their coupling. In Fig.2, the algorithmic steps for the estimation of the AAC are illustrated. Fig. 2A depicts the same trace as the one used in Fig.1A, while Fig.2B and C illustrate the band-pass filtered traces and the related envelopes extracted via Hilbert transform for the low-frequency δ (0.5-4 Hz) and the high-frequency α_1_ (13–20 Hz), respectively. Figure 2D shows the common representation of those envelopes’ traces. The Pearson’s correlation coefficient between every pair of the derived envelope time series was estimated to express the associations between specific frequency pairs. The AAC is computed as follows:

**Figure 2.**
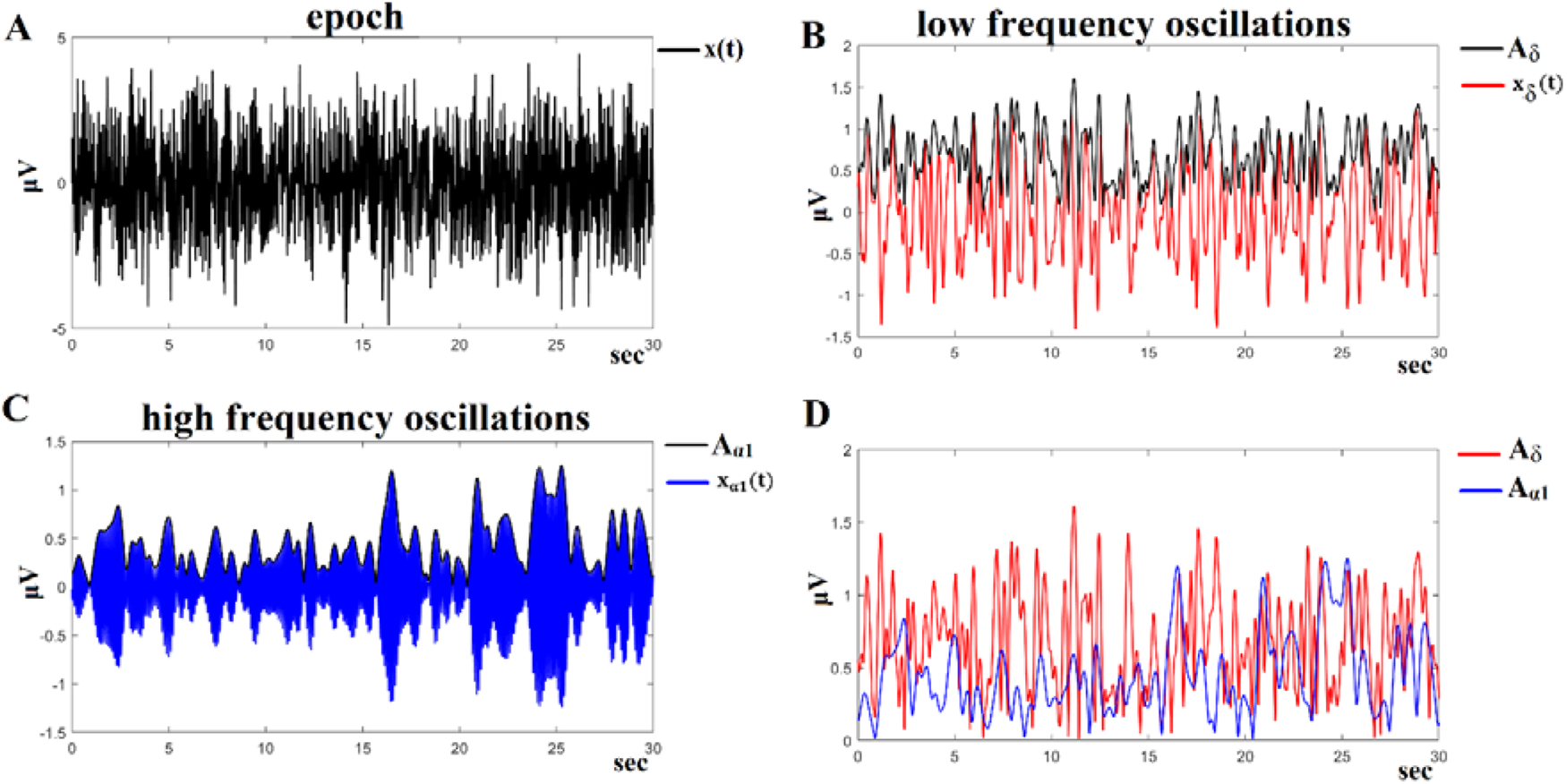
Algorithmic steps for AAC estimation. Illustrating an epoch of 30 sec. length recorded at P4-O2 sensor from the first subject with periodic leg movements during the NREM3 stage (A), we showed the amplitude-to-amplitude coupling between δ and α_1_ rhythms. To estimate δ-α_1_ AAC, the raw signal was band-pass filtered into both (B) a low-frequency δ (0.5–4□Hz) and (C) a high-frequency α_1_ (13–20□Hz) components. The envelopes of those quantities were also extracted via Hilbert transform. Their Hilbert envelopes were demonstrated in conjunction with their band-pass filtered time course (B, C). (D) We then presented into a common plot the envelopes of the bandpass δ (4–8□Hz) and α_1_ (13–20□Hz). The AAC was estimated on these time series using equation 5.

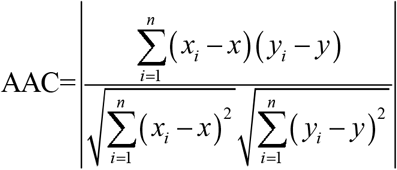

where n is the sample size, *x_i_ y_i_* are the individual sample points indexed with i and 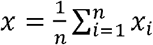 is the sample mean (similarly for y).

Here, we took the absolute values of Pearson’s correlation coefficient. In the present study, we estimated PAC for a total of (7 x 6)/2 = 21 cross-frequency pairs per epoch.

### 2.4 Feature Selection

Our analytic plan extracts an important set of features tailored to CFC. We estimated (CFC) with both estimators across every pair of frequencies and separately for every epoch. Consequently, we averaged the CFC values across epochs of the same sleep stage independently for every subject. The total number of features is: 5 {number of sleep stages} x 2 {number of connectivity estimators} x 21 {possible pair of frequencies} = 210. This set of features was then fed into our Random Forest (RF) model.

### 2.4 Random Forest model and experimental setup

#### 2.5.1 Random Forest

The RF classifier is an ensemble learning method, used extensively in classification and regression tasks [31]. The methodology basically involves the construction of a group (called “forest”) of decision trees as follows: each tree in the “forest” is built on a different training set (called bootstrap sample), drawn randomly from the original training data. A randomly selected variable subset (from the original variable set) is used (during the construction of each decision tree) in order to establish the best split at each node. Once the RF model has been trained and all trees in the forest are constructed, their decisions are combined through majority voting (classification) or averaging (regression). Then, these decisions are used in the prediction of unknown cases.

The RF algorithm provides an internal estimator of a constructed model’s generalization error, called out-of-bag (OOB) error. The estimation of the OOB error involves each decision tree in the model as follows: a portion of the original data cases (around 1/3) do not participate in the training of each tree and instead, are utilized as “test” data, being predicted by the constructed tree. The OOB error estimate is the averaged prediction error for each training case, using only the trees that do not include that case in their bootstrap sample. For more details on the RF algorithm and its underlying notions, we refer for example to [26] and [32].

#### 2.5.2 Experimental setup

In all RF models trained in our experiments, the following parameter setup was used: the number of trees for each model was empirically set based on the OOB error estimate. Following this approach, in all cases, the number of trees was set to 1000. For each RF model and for each node split during the growing of a tree, the number of variables used to determine the best split was set to the default √m (m = total number of features in the dataset). In addition, the RF classifier has its own built-in feature selection method, which was used in order to select the most important features in every RF model. Consequently, only the selected features subsets were utilized for the training of the final RF models. Finally, a 10-fold cross-validation scheme was applied in all experiments for the training of the classifier. It is worth mentioning that the code for our RF-related experiments was written in R, version 3.6.3 (https://www.r-project.org/). Then, the in-house software MATLAB 2019B was used for the estimation of the cross-frequency coupling estimates.

### 2.6 Cyclic Alternating Pattern (CAP): Background and short review

As Cyclic Alternating Pattern (CAP), we characterize sequences of distinct electro-cortical events from the background EEG activity that recur up to 1 min intervals. The existence of CAP may enhance sleep disturbance, instability or both. The appearance of CAP can be spontaneous or in conjunction with sleep disorders, such as e.g. sleep-disordered breathing and periodic leg movement activity. A CAP cycle is composed of a phase A and the following phase B. A CAP sequence is composed of a succession of CAP cycles. CAP sequence has evolved theoretically to encapsulate both sleep maintenance and sleep fragmentation. Phase A is further characterized by subtypes of CAP called A1, A2 and A3. A1 refers to brain’s attempt to maintain sleep related to slow wave activity. Subtypes A2 and A3 CAP constitute an arousal central nervous system that is activated when sleep becomes too unstable or the preservation attempt fails [27]. An index of the quality of sleep is the CAP rate, which is the percentage of the total sum of minutes related to CAP divided by the total sum of NREM sleep. In the present study, we focused on the analysis of non-CAP epochs. For that reason, we estimated the (1 – CAP rate) to report the percentage of NREM time analyses in our study.

## 3. Results

### 3.1 CAP Rates Across Sleep Disorders

Table 3 summarizes the group mean values of (1-CAP rates). It is obvious that (1 - CAP rates) related to the seven sleep disorders deviate from the healthy control group.

**Table 3.** Group averaged (1 - CAP rates)

| **Sleep Disorders** | **Bruxism** | **Insomnia** | **No Pathology (Controls)** | **Narcolepsy** | **Nocturnal frontal lobe epilepsy** | **Periodic leg movements** | **REM behavior disorder** | **Sleep-disordered breathing** |
| --- | --- | --- | --- | --- | --- | --- | --- | --- |
| **(1-CAP Rates) %** | **45.4 ±13.2** | **45.7±11.4** | **51.17±8.61** | **42.39±19.62** | **33.48±10.83** | **29.48±14.72** | **44.72±18.91** | **25.82±17.91** |

### 3.2 Performance of Random Forest Model with CFC Features

Our RF model achieved a high performance of 74% for the P4-O2 EEG sensor, with a macro-average precision of 83.7%, a macro-average recall of 59.1% and a macro-average F-score of 64.9% (Table 4). The rest of the EEG sensors performed worse compared to the optimal scenario: F4-C4 (56%), C4-P4 (56%) and C4-A1 (64%) (see STables 1-3). Table 5 summarizes the confusion matrices of the RF model for the P4-O2 sensor. Finally, the confusion matrices for the other three sensors are presented in the supplementary material of the study (STables 4-6).

**Table 4.** Precision, Recall, F-score and Accuracy performance of the RF model for the P4-O2 EEG sensor

| **Class** | **Precision** | **Recall** | **F-score** |
| --- | --- | --- | --- |
| **brux** | 100% | 50% | 66.7% |
| **ins** | 45.5% | 55.6% | 50% |
| **n** | 87.5% | 87.5% | 87.5% |
| **narco** | 100% | 20% | 33.3% |
| **nfle** | 71.2% | 92.5% | 80.5% |
| **plm** | 80% | 40% | 53.3% |
| **rbd** | 85% | 77.3% | 81% |
| **sdb** | 100% | 50% | 66.7% |
| **Macro-average** | **83.7%** | **59.1%** | **64.9%** |
| **Accuracy** | **74%** |  |  |

**Table 5.** Confusion matrix of the RF model based on the selected features extracted from P4-O2

| **Class** | **brux**<br>**predicted** | **ins**<br>**predicted** | **n**<br>**predicted** | **narco**<br>**predicted** | **nfle**<br>**predicted** | **plm**<br>**predicted** | **rbd**<br>**predicted** | **sdb**<br>**predicted** |
| --- | --- | --- | --- | --- | --- | --- | --- | --- |
| **brux**<br>**real** | 1 | 0 | 0 | 0 | 1 | 0 | 0 | 0 |
| **ins**<br>**real** | 0 | 5 | 0 | 0 | 3 | 0 | 1 | 0 |
| **n**<br>**real** | 0 | 0 | 7 | 0 | 0 | 0 | 1 | 0 |
| **narco**<br>**real** | 0 | 0 | 0 | 1 | 4 | 0 | 0 | 0 |
| **nfle**<br>**real** | 0 | 3 | 0 | 0 | 37 | 0 | 0 | 0 |
| **plm**<br>**real** | 0 | 3 | 0 | 0 | 3 | 4 | 0 | 0 |
| **rbd**<br>**real** | 0 | 0 | 1 | 0 | 4 | 0 | 17 | 0 |
| **sdb**<br>**real** | 0 | 0 | 0 | 0 | 0 | 1 | 1 | 2 |

### 3.3 The Complementarity of PAC-AAC CFC

Figures 3-8 report selected features extracted with both CFC estimators (PAC and AAC) and across sleep stages from P4-O2 sensor. Figure 3 illustrates the seven features related to the AAC across the sleep stages. Figures 4 – 8 demonstrate the twenty-eight selected features from the PAC independently per sleep stage. It is evident that PAC contributes the highest portion of selected features. However, both types encapsulate complementary information under the framework of CFC. The major frequency modulators across sleep stages were δ, θ and α. Interestingly, AAC features were distributed across all sleep stages with the exception of REM.

**Figure 3.**
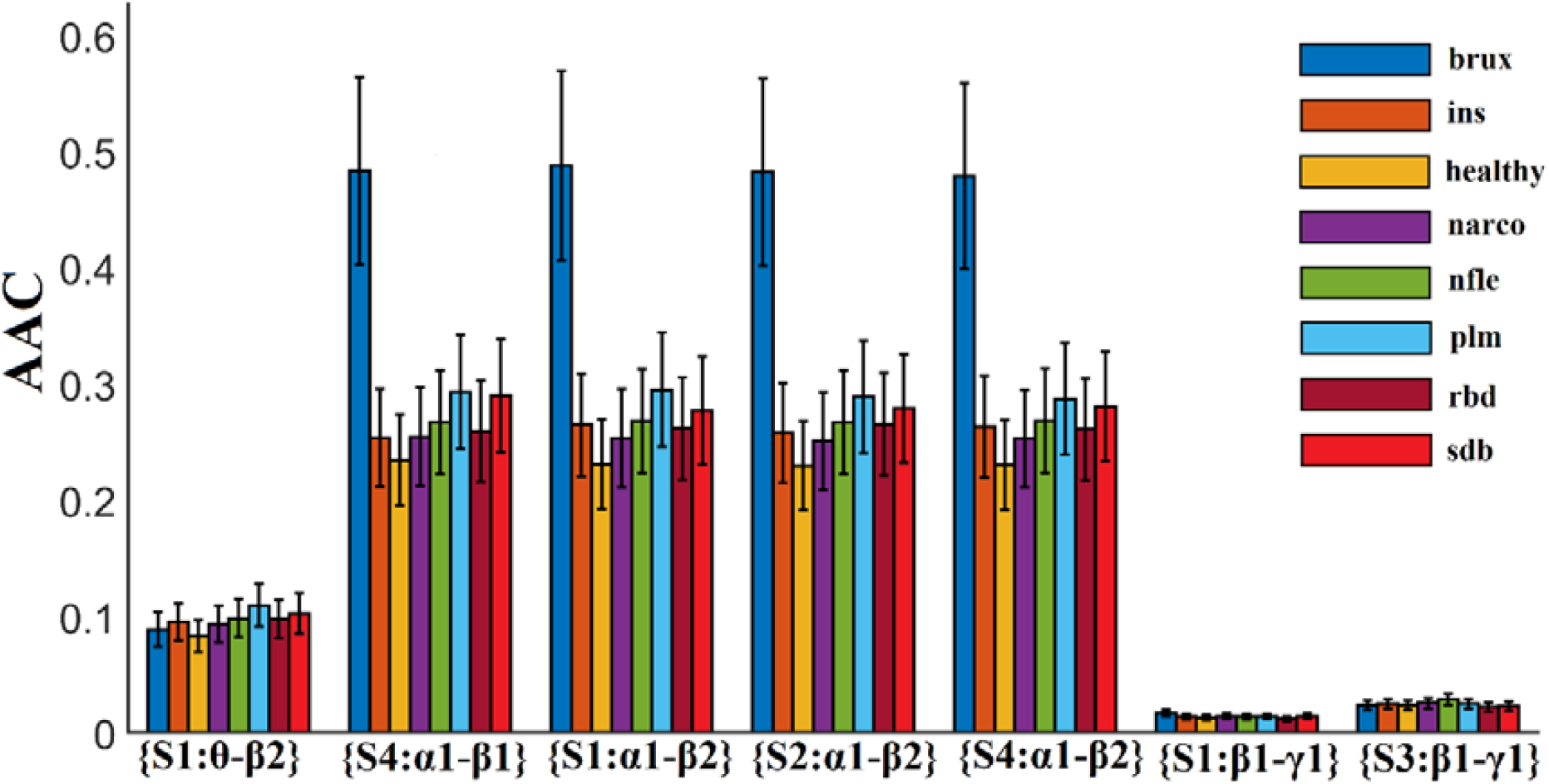
Group-averaged selected AAC values from our random forest model. (S refers to sleep stage) (**Abbreviations of subjects’ pathology for Figures 3 - 8:** Brux – Bruxism, Ins – Insomnia, Healthy – no pathology (controls), Narco – Narcolepsy, Nfle - Nocturnal frontal lobe epilepsy, Plm - Periodic leg movements, Rbd - REM behavior disorder, Sdb - Sleep- disordered breathing)

**Figure 4.**
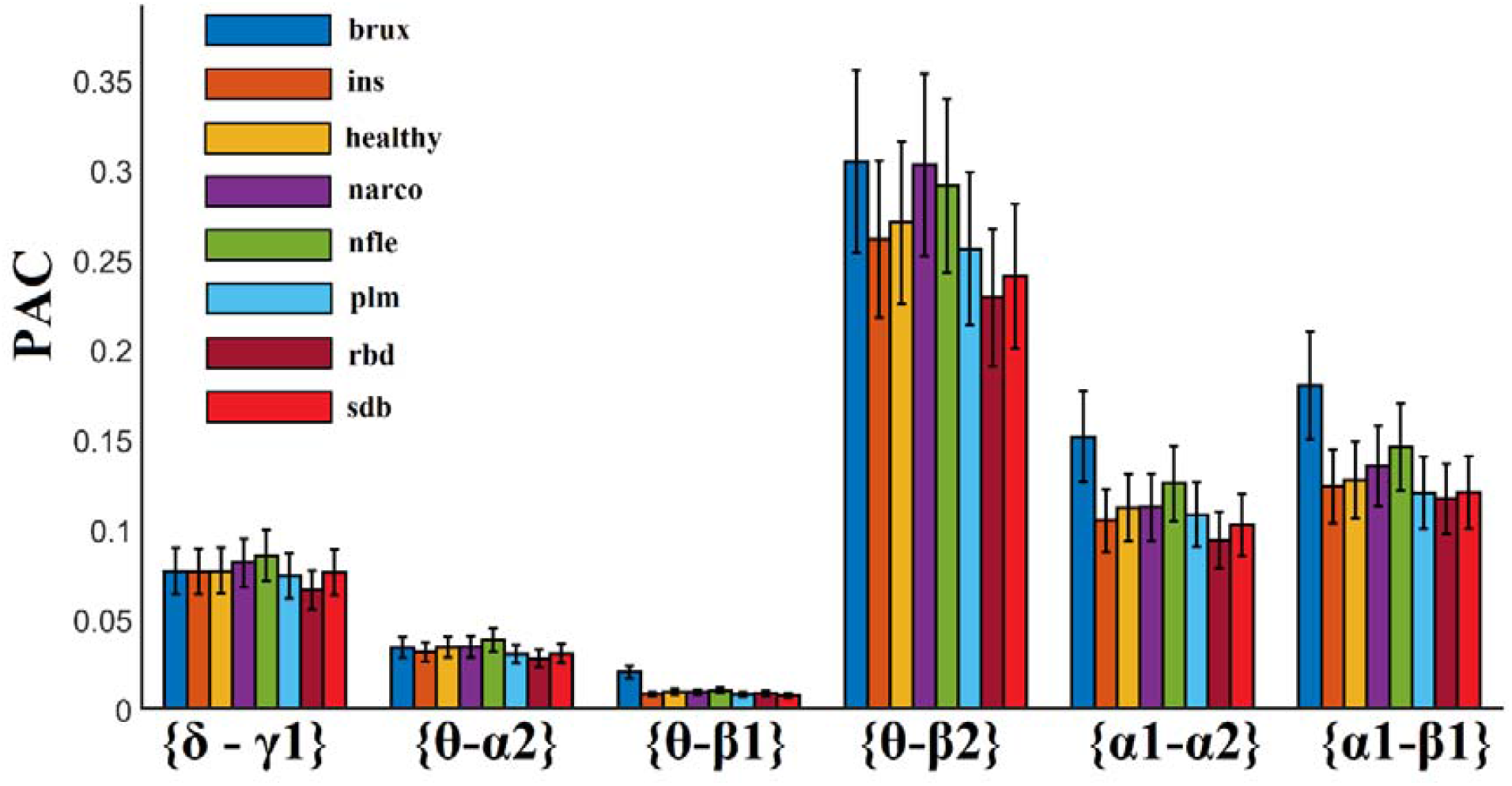
(Sleep stage 1). Group-averaged selected PAC values from our random forest model. (S refers to sleep stage)

**Figure 5.**
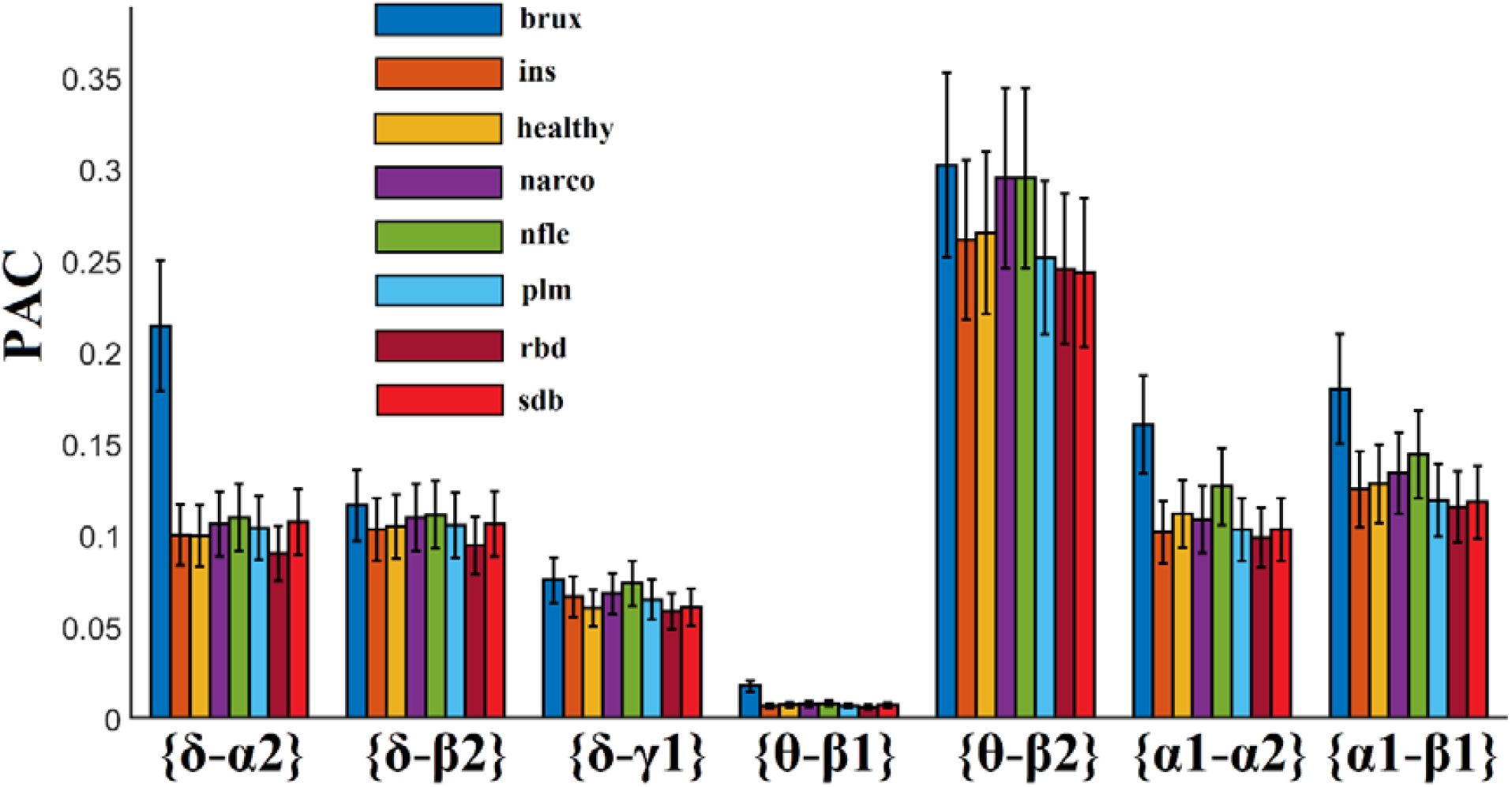
(Sleep stage 2). Group-averaged selected PAC values from our random forest model.

**Figure 6.**
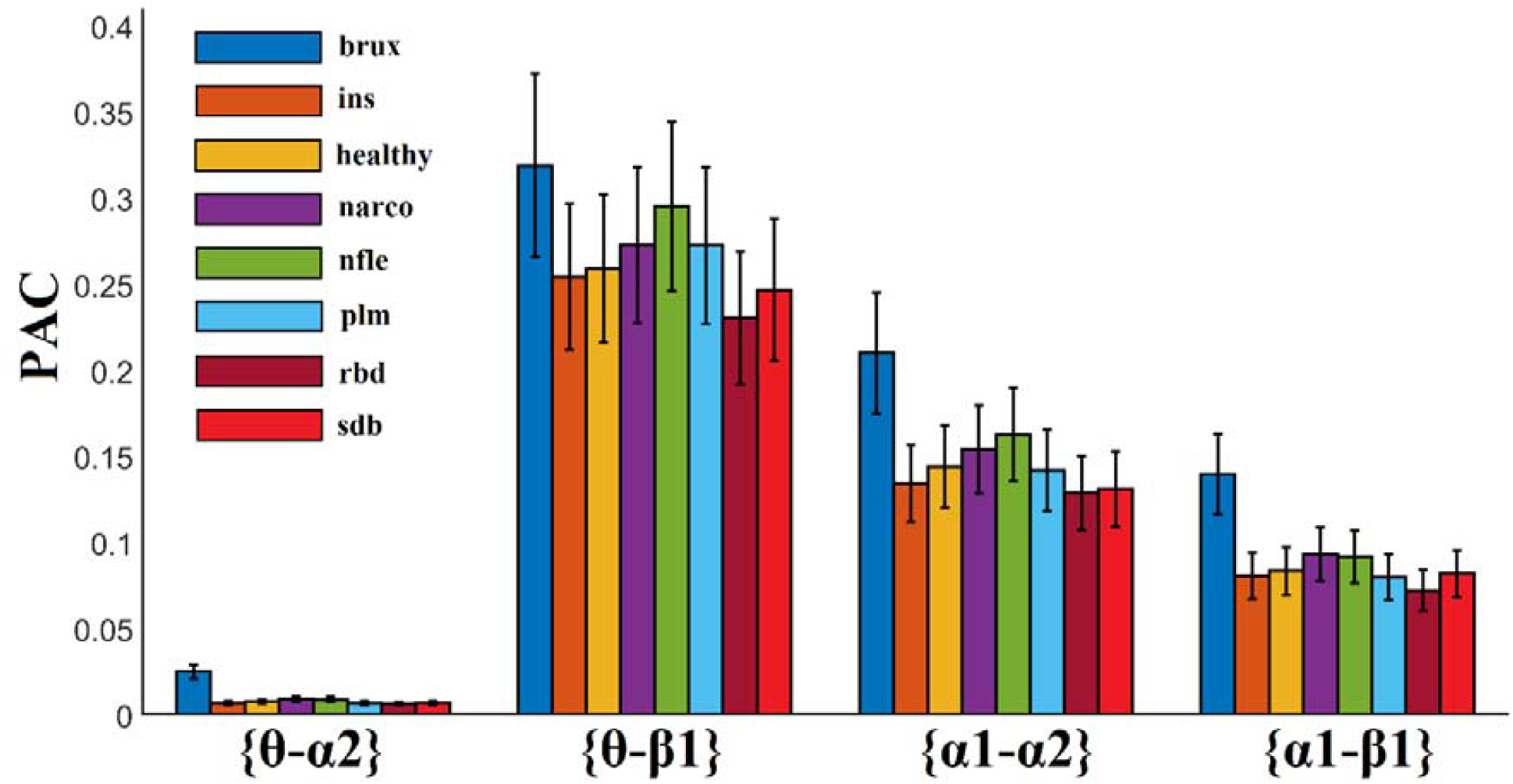
(Sleep stage 3). Group-averaged selected PAC values from our random forest model.

**Figure 7.**
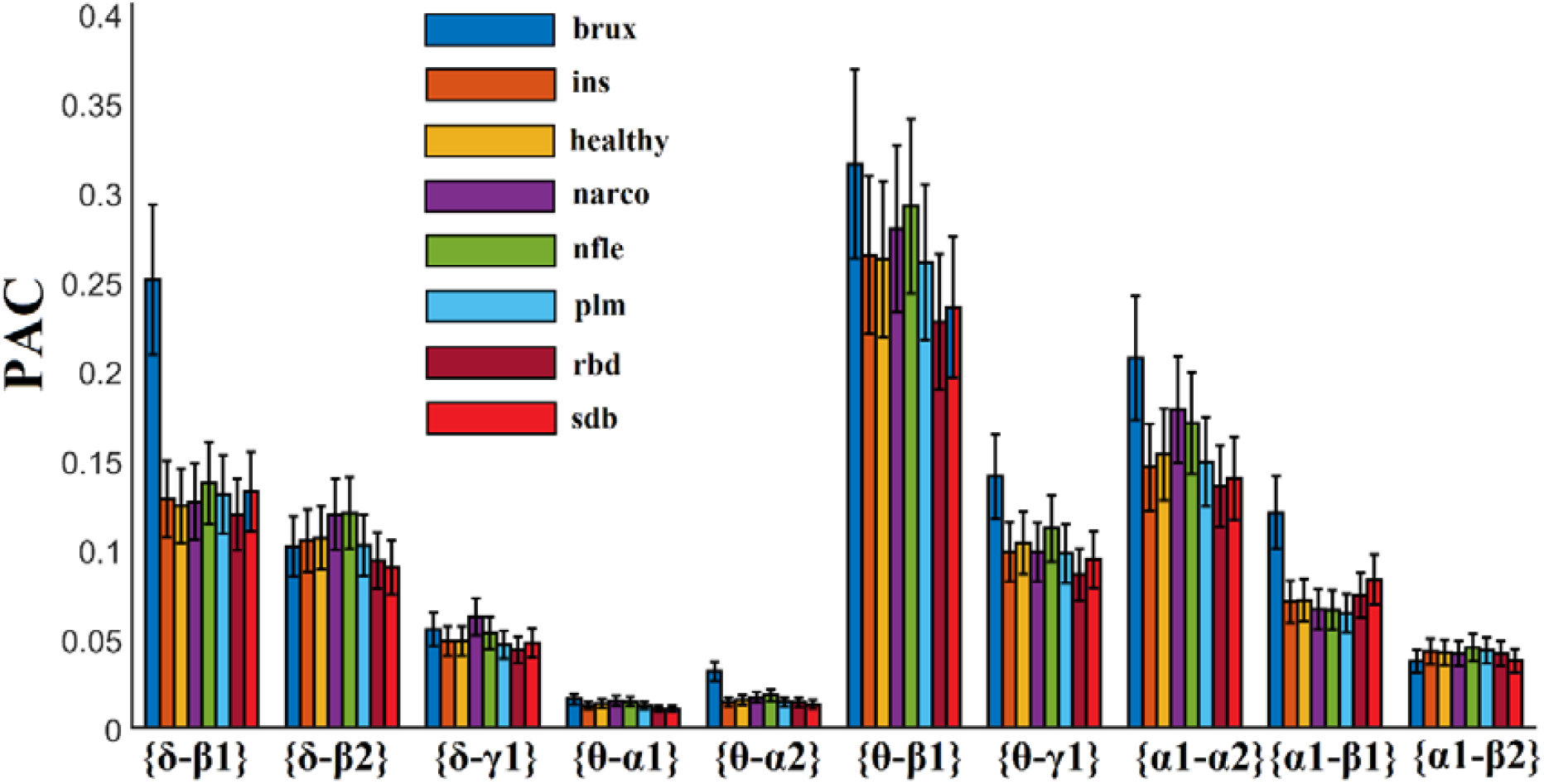
(Sleep stage 4). Group-averaged selected PAC values from our random forest model.

**Figure 8.**
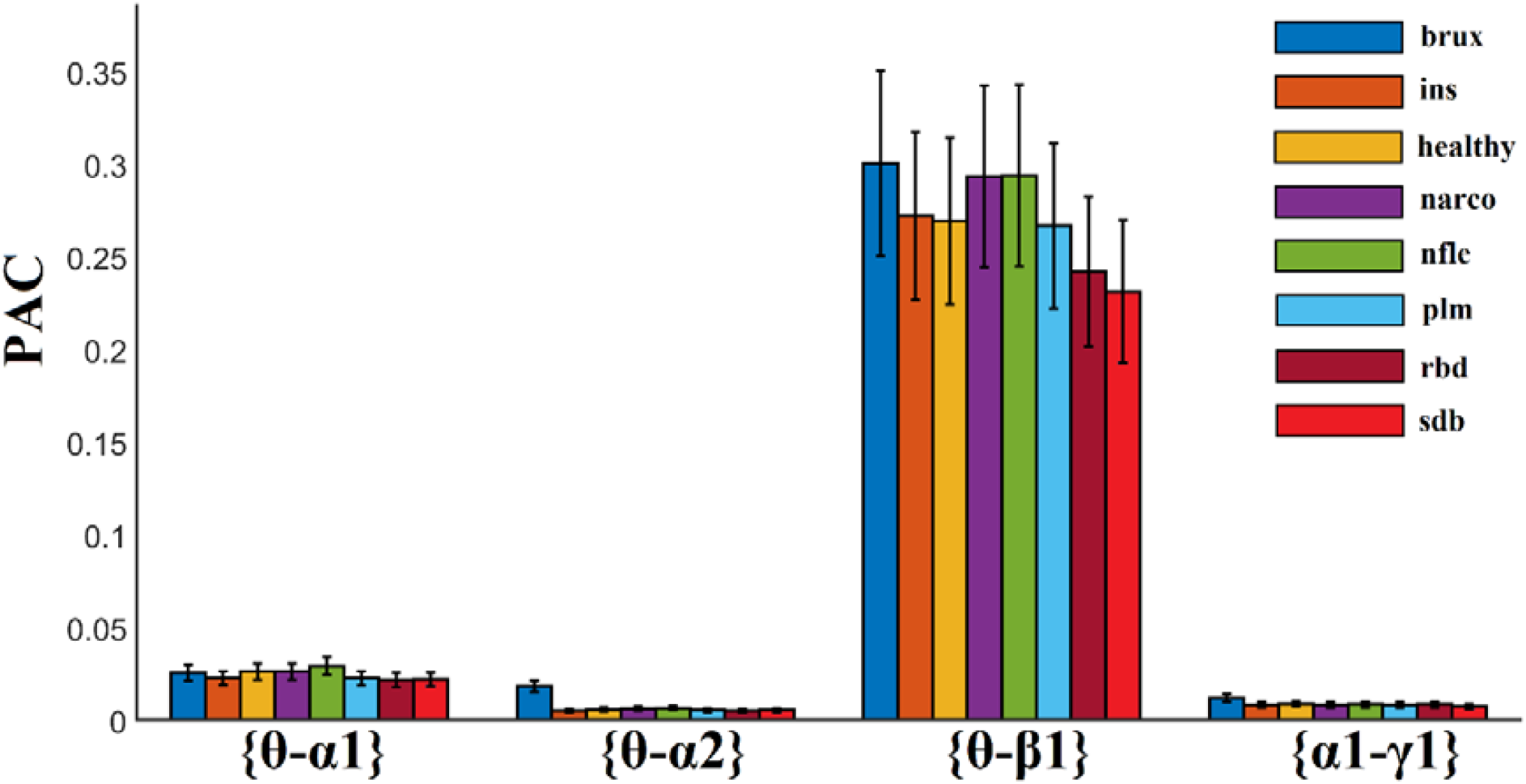
(REM). Group-averaged selected PAC values from our random forest model.

### 3.4 The Importance of Sleep Stage 4

We ran our RF model independently for every sleep stage with the main aim to rank the importance of sleep stages. Our findings ranked sleep stage 4 as the most informative, compared to the other ones for the sleep disorder classification with an accuracy of 62%.

## 4. Discussion

In the present study, we demonstrated the feasibility of complementary CFC estimators computed within multiple EEG sensors across sleep stages and a valuable RF classifier to correctly classify sleep disorders. It is the very first work that attempted to explore how CFC alters on sleep disorders and to classify seven sleep disorders and a healthy control group simultaneously via EEG sleep polysomnographic recordings. We achieved a high classification accuracy of 74% (with macro-average precision: 83.7%, recall: 59.1%, F-score: 64.9%). It should be noted that the most informative common EEG sensor was P4-O2. Our analysis revealed that both types of CFC co-exist during sleep and share complementary information. We also observed similar trends in our recent study, in which we presented a single-sensor automatic sleep stage classification algorithm [13]. Additionally, analyses focusing on single-stage CFC patterns revealed that sleep stage 4 was the most informative in our RF model. Our analysis focused on non-CAP epochs.

The entire methodology of the proposed multiclass sleep disorder detection model can be summarized as follows:

- We kept NREM 3 and NREM 4 as single sleep stages without attempting to merge them [33].
- We adopted an ICA and MODWT (independent component analysis and maximum overlap discrete wavelet transform) to denoise the EEG recordings from eye movements and muscle activity.
- We estimated two different connectivity estimators per frequency pair and for every epoch (one phase-to-amplitude: PAC and one amplitude-to-amplitude: AAC).
- We used a dataset that includes subjects covering seven sleep disorders and a healthy group.
- We analyzed only non-CAP epochs.

Previous studies have attempted to either characterize PAC CFC patterns in sleep disorders or to discriminate sleep disorders using the same dataset. A recent work reported for the very first time that PAC patterns of δ phase modulate α and low-β amplitude in sleep disorders for the three CAP phases [34]. They adopted modulation index as a proper PAC estimator reporting the group-averaged PAC values for the two cross-frequency pairs. This study showed how PAC patterns differed across the three CAP phases at every sleep disorder and also how they differed at every CAP phase across sleep disorders. However, the authors focused only on PAC CFC estimates and only on two cross-frequency pairs without reporting any classification performance. They extracted frequency-dependent signature using an adaptive filtering called empirical mode decomposition (EMD). Another work analysed the same dataset using discrete wavelet transform and related features were extracted [35]. Nonetheless, the authors reported their findings probably by averaging across both epochs and sleep stages with no statistical comparisons between sleep disorders. Moreover, there is no information about the common EEG sensors, from which they estimated and averaged the wavelet-based features. No classification performance has been reported. The third study decomposed every time source into frequency-dependent time courses using EMD, as in the first study [36]. The analysis focused on 20 healthy subjects, 20 REMs, 20 PLMs, and 20 patients with apnea from the same dataset. For every EEG time source, empirical mode decomposition (EMD) returns a data-driven set of intrinsic mode functions called IMFs. Every IMF has a characteristic temporal pattern with a specific frequency profile that can be assigned to well-known brain frequencies. The authors reported that their analysis revealed 9 IMFs, from which high-order statistics have been estimated, like Shannon Entropy, Spectral Entropy, Skewness and Kurtosis and the first order statistic Standard Deviation. Then, they followed a classification approach reporting high performance (93%) between the three sleep disorders and the healthy control group for features related to IMF8. However, they didn’t report how and where the whole methodology was applied, which sensor they analysed, as well as how they managed the different epochs and the sleep stages. None of the aforementioned studies denoised the original EEG traces.

Brain frequencies can couple between each other via different but complementary mechanisms. The basic CFC mechanisms are phase-to-amplitude (PAC) and amplitude-to-amplitude envelope correlation (AAC) [37, 38]. The first CFC mechanism, called PAC, quantifies how the phase of the slow frequency modulates the amplitude of the higher frequency. The explanation of the existence of such a mechanism and how slow oscillations can couple faster brain frequencies in many brain areas is on the conduction velocities of cortical neurons. Slower frequencies activate more neurons in large brain volumes associated with large changes of membrane potential [39]. This practically means that in longer temporal windows, a larger portion of spikes linked to upstream neurons can co-exist [40]. There are evidences that PAC exists between every possible pair of mammalian brain frequencies from 0.025 Hz up to 500 Hz [41] and in sleep [13, 42]. A complementary CFC mechanism to PAC is the AAC, which captures the temporal covariation between the envelopes of the slower and the faster frequencies [43]. This approach is temporally less precise, but important to capture CFC mechanisms in many cases and especially in sleep [13].

In our study, we identified that the P4-O2 EEG sensor provided the highest accuracy among the four common EEG sensors. NREM 4 was also the most informative sleep stage following a classification process independently for every sleep stage. Our analysis supported the notion of analysing NREM 3 and 4 separately. The conclusion is that the combination of PAC and AAC in NREM 4 estimated over parieto-occipital brain area provided the most powerful sleep disorder detection model so far in the literature. It is known that consolidation processes rely on the interactions between characteristic brain oscillatory activity of NREM sleep: slow oscillations, spindles and ripples [44]. A recent study measured intracranial EEG activity during sleep from epileptic patients. They reported PAC between 0.5–1.25 Hz (slow oscillation range) and 12–16 Hz (spindle range) within the Cz sensor. Their findings were also evaluated from intracranial recordings from hippocampus, providing a link of PAC estimates between EEG sensor activity and intracranial findings. PAC estimates revealed the hierarchical role of slow oscillations, spindles and ripples during NREM [44,45]. Both studies [44,45] evaluated the existence of PAC as a brain connectivity pathway within specific brain areas while they validated a clear link between sensor and source brain activity.

To the best of the authors’ knowledge, the current study provided the best performance of a sleep disorder detection model in the literature. Our analysis involved the estimation of two CFC mechanisms among many frequency pairs within four common EEG sensors and across five sleep stages. The RF model served as a natural multiclass classifier and not as a one-versus-all classification approach. This work is another support of CFC existence during sleep [13]. We further proved that sleep disorders altered both types of CFC mechanisms while PAC estimator was more informative compared to AAC, supplying our RF model with more features. Figure 3 illustrates the seven features related to the AAC across the sleep stages, while Figures 4 – 8 demonstrate the twenty-eight selected features from the PAC independently per sleep stage. In the future, we will adopt the same strategy tailored to the three CAP phases in conjunction with complementary CAP related features like the CAP rates, which is the ratio of the durations in secs between NREM CAP sleep and total NREM sleep [46].

This study provides evidence that two basic CFC are sensitive to detect alterations of nested oscillations across sleep disorders in various sleep stages. We reported important findings of the existence of a multiplex nature of brain oscillations in sleep that is altered due to specific disorders. Additionally, PAC and AAC showed that they capture complementary information in relation to CFC mechanisms. Here, our study focused on the estimation of CFC within sensors and only in non-CAP epochs. Further analysis is important to explore CFC patterns between EEG sensors and also in CAP epochs. We aim to explore the aforementioned scenarios in a complementary study with the same dataset. Additionally, the classification performance of CFC patterns should be compared also with various CAP related features to further validate their importance in an accurate sleep disorder detection model. Moreover, a relationship between CFC patterns and CAP rates should be reported.

One of the limitations of this study comes from the restrictions of the current database in terms of the imbalance of participants across sleep disorders. EEG features and especially brain connectivity are sensitive to sex and age parameters [47]. CFC has not yet demonstrated its sensitivity on age and sex during sleep. Carefully reading the confusion matrix tabulated in Table 4, one can make the conclusion that the misclassified subjects varied across age and also sex. Age range of misclassified PLM, RBD and Insomnia subjects range from 48 up to 70 years. We assume that a database of subjects with no sleep pathology across the lifespan would have eliminated any doubt about the sensitivity of our model over a misbalanced dataset with age. More subjects are needed to further cross-validate our model, while a high-density EEG sleep recording of subjects with sleep disorders would support the source localization of brain activity in virtual brain areas to explain current findings with CFC.

With respect to the imbalanced nature of the dataset and the selection of the classification algorithm, we would like to note that the decision to employ Random Forest (RF) in order to build the classification model in our study was based on the fact that after preliminary observations, RF outperformed all other algorithms that were used in our initial experiments. In addition, RF is generally robust to overfitting and is frequently preferred in classification problems involving highly imbalanced datasets.

It is important to mention here that our analysis is based on the hypnograms created by experts and also on the distinction between CAP and non-CAP epochs. Data-driven characterization of our model includes the artefact correction, the estimation of CFC-based features and also the classification approach with RF. To make this approach fully automated, we have to apply an automatic sleep stage classification algorithm [26] and an automatic characterization of the CAP epochs.

An important sub-class of sleep disorders is sleep-related movement disorders. Within this category, the following clinical conditions are included: periodic limb movement disorder (PLMD), restless leg syndrome (RLS), sleep-related leg cramps, and bruxism [48]. In clinical practice, an expert visualizes EMG activity of the anterior tibialis to detect epochs of limb movements. If EMG activity overcomes the threshold of 8 microvolts at rest with a duration between 0.5 and 10 seconds, then this epoch is characterized as limb movement [48]. According to ICSD-3 of the American Academy of Sleep Medicine, the diagnostic criteria for PLMS are: the detection of more than 15 periodic limb movements per hour in adults and more than 5 periodic limb movements per hour in children that cause in both cases strong sleep problems [49]. In the present study, we aimed to apply a common analytic framework across sleep disorders using EEG-based characteristics. It is well-known that periodic limb movements in sleep alter EEG activity via the arousal response, going from autonomous activation to alterations of EEG activity across the frequency range [50].

We intend in a future study to investigate CFC patterns in CAP epochs and also before and during the occurrence of K-complexes and sleep spindles. The comparison of the potential findings of such a study with findings presented here on non-CAP epochs and especially their complementarity will shed light on the importance of EEG modality and CFC patterns, on the understanding of sleeps disorders, as well as on their correct classification.

## 5. Conclusion

In the present study, we reported for the very first time a machine learning approach of an automatic classification system of sleep disorders, based on EEG modality and complementary cross-frequency system. The whole analysis focused on non-CAP epochs from NREM stages and also on REM stage. Our system succeeded a performance of 74% to correctly classify a subject across seven sleep disorders and a healthy control group. Further exploratory studies are needed, including the repetition of the same analysis also onCAP epochs including also alternative CFC estimators. It would be interesting to also compare the importance of EEG-based analysis in comparison to CAP-related features and also during specific epiphenomenon, such as K-complexes and sleep spindles.

## Data Availability

https://physionet.org/content/capslpdb/1.0.0/

## Author Contributions

**Stavros I. Dimitriadis:** conceptualization, methodology, formal analysis, software, writing (original draft).

**Christos I. Salis:** data curation, performed the analysis, writing (reviewing and editing).

**Dimitris Liparas:** data curation, performed the machine learning, writing (reviewing and editing).

Every author read and approved the final version of the manuscript.

## Funding

SID was supported by MRC grant MR/K004360/1 (Behavioral and Neurophysiological Effects of Schizophrenia Risk Genes: A Multi-locus, Pathway Based Approach) and by a MARIE-CURIE COFUND EU-UK Research Fellowship.

## Conflict of Interest

The authors declare that the research was conducted in the absence of any commercial or financial relationships that could be construed as a potential conflict of interest.

## Acknowledgments

We would like to acknowledge researchers, staff and technicians that made this dataset available to the public. Special thanks also to staff and scientists that maintain physionet database active.

## Disclosure Statement

- Financial Disclosure: none.
- Non-financial Disclosure: non

### Abbreviations

PLV: Phase locking value
iPLV: Imaginary part of PLV
CFC: Cross-frequency coupling
PAC: phase-amplitude coupling
AAC: amplitude-amplitude coupling
EMD: empirical mode decomposition
IMF: intrinsic mode functions
CAP: cyclic alternating pattern

## Notes

### Competing Interest Statement

The authors have declared no competing interest.

### Clinical Trial

The study involves the analysis of preexisting datasets that have been downloaded from:
https://physionet.org/content/capslpdb/1.0.0/

### Clinical Protocols

https://physionet.org/content/capslpdb/1.0.0/

### Author Declarations

Everything is described in the original articles. https://physionet.org/content/capslpdb/1.0.0/

### Summary of Updates

We revised the parts of the document.

## References

1. Younes M. The case for using digital EEG analysis in clinical sleep medicine. SSP. 2017; 1(1): 2

2. Solms, M. Dreaming and REM sleep are controlled by different brain mechanisms. Behav. Brain Sci. 2000; 23(6): 843–850.

3. Zee P. C., Vitiello MV. Circadian rhythm sleep disorder: irregular sleep wake rhythm type. Sleep Med. Clin. 2009; 4(2): 213–218.

4. Thorpy M. J. Classification of sleep disorders. Neurotherapeutics. (2012); 9(4): 687–701.

5. National Institutes of Health. National Institutes of Health Sleep Disorders Research Plan.Last accessed May 30, 2020 https://www.nhlbi.nih.gov/files/docs/ncsdr/201101011NationalSleepDisordersResearchPlanDHHSPublication11-7820.pdf.

6. Sleep Disorder Classifications - SleepHealth, SleepHealth, 2020. Accessed: 31-May-2020 https://www.sleephealth.org/sleep-health/sleep-disorder-classifications/.

7. Davis H., Davis P. A., Loomis A. L., Harvey E. N., Hobart G. Human brain potentials during the onset of sleep. J. Neurophysiol. 1938; 1(1): 24–38.

8. Canolty R. T., Knight R. T. The functional role of cross-frequency coupling. Trends Cogn. Sci. 2010; 14 (11): 506–515. doi: 10.1016/j.tics.2010.09.001.

9. Haustein W., Pilcher J., Klink J., Schulz H. Automatic analysis overcomes limitations of sleep stage scoring. Electroencephalogr. Clin. Neurophysiol. 1986; 64(4): 364–374. doi: 10.1016/0013-4694(86)90161-6.

10. Al-Salman W., Li Y., Wen P. Detection of EEG k-complexes using fractal dimension of time frequency images technique coupled with undirected graph features. Front. Neuroinform. 2019; 13: 45. https://doi.org/10.3389/fninf.2019.00045

11. Jenni O. G., Carskadon M. A. Spectral analysis of the sleep electroencephalogram during adolescence. Sleep. 2004; 27 (4): 774–783.

12. Kurth S., Ringli M., Geiger A., LeBourgeois M., Jenni O. G., Huber R. Mapping of cortical activity in the first two decades of life: a high-density sleep electroencephalogram study. J Neurosci. 2010; 30 (40): 13211–13219.

13. Dimitriadis S. I., Salis C., Linden D. A novel, fast and efficient single-sensor automatic sleep-stage classification based on complementary cross-frequency coupling estimates. Clinical Neurophysiology. 2018; 129 (4): 815–828

14. Dimitriadis S. I., Laskaris N. A., Bitzidou M. P., Tarnanas I., Tsolaki M.. A novel biomarker of amnestic MCI based on dynamic Cross-Frequency Coupling patterns during cognitive brain responses. Front. Neurosci. 2015; 9: 350. doi: 10.3389/fnins.2015.00350

15. Antonakakis, M., Dimitriadis, S. I., Zervakis, M., Rezaie, R., Babajani-Feremi, A., Micheloyannis, S., et al. Detecting mild traumatic brain injury from resting-state MEG recordings based on Cross-frequency Interactions. Int. J. Psychophysiol. 2016; 102: 1–11.

16. Belluscio M. A., Mizuseki K., Schmidt R., Kempter R., Buzsáki G. Cross-frequency phase-phase coupling between theta and gamma oscillations in the hippocampus. J. Neurosci. 2012; 32 (2): 423–435.

17. Jensen O., Colgin L. L. Cross-frequency coupling between neuronal oscillations. Trends Cogn. Sci. 2007; 11(7): 267–269.

18. Axmacher, N. et al. Cross-frequency coupling supports multi-item working memory in the human hippocampus. (2010) Proc. Natl. Acad.Sci. 107, 3228–3233.

19. Schnitzler A and J. Gross, Normal and pathological oscillatory communication in the brain. 2005; Nat. Rev. Neurosci., 6 (4): 285–296.

20. Canolty R. T., Edwards E., Dalal S. S., Soltani M., Nagarajan S. S., Kirsch H. E., Berger M. S., Barbaro N. M., Knight R. T. High gamma power is phase-locked to theta oscillations in human neocortex, Science, 2006; 313 (5793): 1626–1628.

21. Scheffzük C., Kukushka V. I., Vyssotski A. L., Draguhn A., Tort A. B. L., Brankaφk J., Selective coupling between theta phase and neocortical fast gamma oscillations during REM-sleep in mice, PloS One, 2011; 6 (12): e28489.

22. Szczepanski S. M., Crone N. E., Kuperman R. A., Auguste K. I., Parvizi J., Knight R. T., Dynamic changes in phase-amplitude coupling facilitate spatial attention control in fronto-parietal cortex. PLoS Biol., 2014; 12(8): e1001936.

23. Palva J. M., Palva S., Kaila K. Phase synchrony among neuronal oscillations in the human cortex. J. Neurosci. 2005; 25 (15): 3962–3972.

24. Dimitriadis S. I., Sun Y. U., Kwok K., Laskaris N. A., Thakor N., Bezerianos A. Cognitive workload assessment based on the tensorial treatment of EEG estimates of cross-frequency phase interactions. Ann. Biomed. Eng. 2015; 43 (4): 977–989. doi: 10.1007/s10439-014-1143-0

25. Shirvalkar P. R., Rapp P. R., Shapiro M. L. Bidirectional changes to hippocampal thetagamma comodulation predict memory for recent spatial episodes, Proc. Natl. Acad. Sci., 2010; 107 (15): 7054–7059

26. Dimitriadis SI, Liparas D, Tsolaki MN. Alzheimer’s Disease Neuroimaging Initiative. Random forest feature selection, fusion and ensemble strategy: Combining multiple morphological MRI measures to discriminate among healhy elderly, MCI, cMCI and alzheimer’s disease patients: From the alzheimer’s disease neuroimaging initiative (ADNI) database. J Neurosci Methods. 2018; 302: 14–23.

27. Terzano, M. G. et al. Atlas, rules, and recording techniques for the scoring of cyclic alternating pattern (CAP) in human sleep. Sleep Med. 2001; 2 (6): 537–553.

28. Goldberger A. L. et al. PhysioBank, PhysioToolkit, and PhysioNet: Components of a new research resource for complex physiologic signals. Circulation, 2000; 101 (23): e215–e220.

29. Wolpert E. A. A manual of standardized terminology, techniques and scoring system for sleep stages of human subjects. Arch. Gen. Psychiatry. 1969; 20 (2): 246–247.

30. Voytek B., Canolty R.T., Shestyuk A., Crone N.E., Parvizi J., Knight R.T. Shifts in gamma phase-amplitude coupling frequency from theta to alpha over posterior cortex during visual tasks. Front. Hum. Neurosci. 2010; 4: 19

31. Breiman, L. Random forests. Machine learning, 2001; 45 (1): 5–32.

32. Liparas D., HaCohen-Kerner Y., Moumtzidou A., Vrochidis S., Kompatsiaris I. News articles classification using random forests and weighted multimodal features. In Information Retrieval Facility Conference, 2014; 63–75. Springer, Cham.

33. Tsinalis O, Matthews PM, Guo Y. Automatic sleep stage scoring using time frequency analysis and stacked sparse autoencoders. Ann. Biomed. Eng. 2016; 44(5): 1587–1597.

34. Yeh C. H., Shi W. Identifying phase-amplitude coupling in cyclic alternating pattern using masking signals, Sci. Rep., 2018; 8(1): 2649.

35. Rao T., Vishwanath D. D. Detecting sleep disorders based on EEG signals by using discrete wavelet transform. In: 2014 International conference on green computing communication and electrical engineering (ICGCCEE). 2014: 1–4, Coimbatore, India.

36. Islam M.R., Rahim M.A., Akter H., Kabir R., Shin J.: Optimal IMF selection of EMD for sleep disorder diagnosis using EEG signals. In: Proceeding of the 3rd International Conference on Applications in Information Technology, 2018: 96–101, AizuWakamatsu, Japan.

37. Buzsaki G, Draguhn A. Neuronal oscillations in cortical networks. Science, 2004; 304: 1926–1929. https://doi.org/10.1126/science.1099745.

38. Buzsáki G, Watson BO. Brain rhythms and neural syntax: implications for efficient coding of cognitive content and neuropsychiatric disease. Dialogues Clin Neurosci 2012; 14 (4): 345–367.

39. Von Stein A, Sarnthein J. Different frequencies for different scales of cortical integration: from local gamma to long range alpha/theta synchronization. Int J Psychophysiol 2000; 38 (3): 301–313. https://doi.org/10.1016/S0167-8760(00)00172-0.

40. Quilichini P, Sirota A, Buzsaki G. Intrinsic circuit organization and theta-gamma oscillation dynamics in the entorhinal cortex of the rat. J Neurosci, 2010; 30 (33): 11128– 11142.

41. Sirota A, Csicsvari J, Buhl D, Buzsaki G. Communication between neocortex and hippocampus during sleep in rodents. Proc Natl Acad Sci USA. 100 (4) 2003; 100 (4): 2065– 2069.

42. Amiri M, Frauscher B, Gotman J. Phase-amplitude coupling is elevated in deep sleep and in the onset zone of focal epileptic seizures. Front Hum Neurosci 2016; 10: 387 https://doi.org/10.3389/fnhum.2016.00387.

43. Bruns A., Eckhorn R., Gail A., Brinksmeyer H. J., Schanze T. Directional coupling of gamma-envelopes and theta-signals between separate neuronal populations in human and monkey visual cortex. Soc. Neurosci. Abstr., 2001; 27: 36.

44. Staresina B. P., Bergmann T. O., Bonnefond M., Van der Meij R., Ole J., Deuker L., et al. Hierarchical nesting of slow oscillations, spindles and ripples in the human hippocampus during sleep. Nat Neurosci, 2015; 18: 1679–1686.

45. Andrillon T., Nir Y., Staba R. J., Ferrarelli F., Cirelli C., Tononi G., Fried I. Sleep spindles in humans: Insights from intracranial EEG and unit recordings. J. Neurosci. 2011; 31 (49): 17821–17834.

46. Terzano M. G., Mancia D., Salati M. R., Costani G., Decembrino A., Parrino L. The cyclic alternating pattern as a physiologic component of normal NREM sleep. Sleep, 1985; 8 (2): 137–145.

47. Ujma PP, Konrad BN, Simor P, Gombos F, Körmendi J, Steiger A, Dresler M, Bódizs R. Sleep EEG functional connectivity varies with age and sex, but not general intelligence. Neurobiol Aging. 2019 Jun;78:87–97.

48. Joseph V, Nagalli S. Periodic Limb Movement Disorder. 2020 Jul 15. In: StatPearls [Internet]. Treasure Island (FL): StatPearls Publishing; 2021 Jan–. PMID: 32809562.

49. Stefani A, Högl B. Diagnostic Criteria, Differential Diagnosis, and Treatment of Minor Motor Activity and Less Well-Known Movement Disorders of Sleep. Curr Treat Options Neurol. 2019 Jan 19;21(1):1

50. Sforza E, Nicolas A, Lavigne G, Gosselin A, Petit D, Montplaisir J. EEG and cardiac activation during periodic leg movements in sleep. Support for a hierarchy of arousal responses. Neurology 1999a;52:786–91

